# Smoking, distress and COVID-19 in England: cross-sectional population surveys from 2016 to 2020

**DOI:** 10.1101/2020.12.07.20245514

**Authors:** Loren Kock, Jamie Brown, Lion Shahab, Graham Moore, Marie Horton, Leonie Brose

## Abstract

**Background:** Changes in the prevalence of mental health problems among smokers due to the COVID-19 pandemic in England have important implications for existing health inequalities. This study examined the prevalence of psychological distress among smokers following the onset of the pandemic compared with previous years.

**Methods:** Cross-sectional data were used from a representative survey of smokers (18+) in England (n = 2,927) during four months (April to July) in 2016, 2017 and 2020. Adjusted logistic regressions estimated the associations between past-month psychological distress across two time periods (2016/17 and 2020), and age. Weighted proportions, chi-squared statistics and stratified logistic regression models were used to compare the distributions of minimal, moderate and severe distress, respectively, within socio-demographic and smoking characteristic categories in 2016/17 and 2020.

**Results:** The prevalence of moderate and severe distress among past-year smokers was higher in 2020 (moderate: 28.79%, 95%CI 26.11-31.60; OR=2.08, 95%CI 1.34-3.25; severe: 11.04%, 9.30-13.12; OR=2.16, 1.13-4.07) than in 2016/17 (moderate: 20.66%, 19.02-22.43; severe: 8.23%, 7.16-9.47). While there was no overall evidence of an interaction between time period and age, young (16-24 years) and middle-age groups (45-54 years) may have experienced greater increases in moderate and older age groups (65+ years) in severe distress from 2016/17 to 2020. There were also increases in 2020 of moderate distress among those from more disadvantaged social grades and of both moderate and severe distress among women and those with low cigarette addiction.

**Conclusions:** Between April-July 2016/17 and April-July 2020 in England there were increases in both moderate and severe distress among smokers. The distribution of distress differed between 2016/17 and 2020 and represents a widening of established inequalities, with increases in distress among socio-economically disadvantaged groups, women and diverging age groups.

**What this paper adds**

- Surveys in the UK indicate that there has been a deterioration in mental health of the general population since the onset of the COVID-19 pandemic.
- Smoking is strongly associated with poor mental health, and a deterioration in mental health among smokers has potentially damaging consequences for existing health inequalities in the UK.
- Our findings using data from a large population-based sample of adults in England show that between 2016/17 and 2020 there were increases in moderate and severe psychological distress among smokers.

## INTRODUCTION

Between 2014 and 2015 the prevalence of adult smoking in England was estimated to be 16.4%.^1^ Across the same period smoking prevalence was higher among those with anxiety or depression (28.0%), a long-term mental health condition (34.0%) and serious mental illness (40.5%).^1^ Those with a mental health condition are more likely to be more dependent smokers and to have greater difficulty in remaining abstinent after quitting, despite greater desire to quit compared with the general population.^2^ These differences in smoking may account for up to two thirds of the inequality in life expectancy between those living with and without a mental health condition.^3^ This study aimed to examine the prevalence of distress among smokers following the onset of the COVID-19 pandemic in England compared with previous years.

Serious psychological distress is defined as mental health problems that are associated with impairment in social or occupational functioning and require treatment.^4^ Psychological distress is more common among smokers and is negatively associated with quit success and abstinence.^5,6^ The relationship between smoking and distress may be explained by common risk factors related to socio-economic position^7^, but research also suggests potential bidirectionality^8^. Individuals may be motivated to smoke to alleviate symptoms of distress, and there is evidence that smoking may itself directly increase the risk of distress.^9,10^

Between 2016-2017 in England, 24.3% and 9.7% of past-year smokers indicated moderate and serious past-month distress, respectively.^11^ Also, those with an indication of a mental health problem were more dependent on cigarettes but more likely to be motivated and have recently attempted to quit.

The COVID-19 pandemic and resulting government ‘lockdown’ measures were associated with a deterioration of mental health in the UK compared with pre-COVID-19 trends, with specific burden among women and young adults.^12,13–15^ Research has suggested that following the March 2020 government restrictions, smokers were more likely to try and quit, and rates of smoking cessation were higher.^16^ However, a deterioration in mental health among smokers may negatively impact quitting behaviour given that smokers with distress have been found to be less likely to quit and remain abstinent.^5,6^

Our previous research has highlighted an age gradient in distress among smokers, with younger groups reporting higher levels of distress compared with older age groups.^11^ However, considering the sharp positive age gradient in the risk of death from COVID-19^17^, deterioration in mental health during the pandemic may be more pronounced among older age groups.

An increase in the prevalence of distress among smokers during the COVID-19 pandemic in England could potentially widen existing health inequalities. Monitoring levels of distress among smokers is important to highlight unmet need for mental health and smoking cessation support in general and also during current and potential future respiratory disease epidemics. Using Smoking Toolkit Study (STS) data the aims of this study were to i) examine the prevalence of psychological distress among past-year smokers during April-July 2020 compared with the same monthly time period (April-July) in 2016-2017 (the previous time distress was assessed in the STS) and ii) examine the distribution of distress within sociodemographic and smoking characteristic categories of past-year smokers during April-July 2020 and April-July 2016-2017.

## METHODS

### Study design

Data were drawn from the Smoking Toolkit Study (STS), a monthly repeated cross-sectional survey of a representative sample of adults in England.^18^ The dataset for the primary analysis consisted of four months of STS data from April to July in each of the years 2016, 2017 and 2020. Respondents were age 18 years or older.

The STS uses a hybrid of random location and quota sampling to select a new sample of approximately 1,700 adults each month. Locations are randomly selected from around 170,000 output areas in England stratified by geodemographic characteristics. Interviews are performed with one household member until quotas based on factors influencing the probability of being at home (e.g. gender, age, working status) are fulfilled. Comparisons with other national surveys show that the STS recruits a representative sample of the population in England.^18^ Data are usually collected monthly through face-to-face computer assisted interviews. However, due to the COVID-19 pandemic, from March 2020 data were collected via telephone only. Diagnostic analyses have suggested it is reasonable to compare data from before and after the lockdown, despite the change in data collection method.^16^

Ethical approval for the STS is granted by the UCL Ethics Committee (ID 0498/001; ID: 2808/005). The Strengthening the Reporting of Observational Studies in Epidemiology (STROBE) reporting guideline were used in the design and reporting of this study.^19^

### Dependent variables, independent variables and covariates

The primary outcome of this study was the prevalence of psychological distress among past-year smokers. This was derived using the following measures.

#### Mental health (Past-month psychological distress)

Past**-**month distress was measured using the K6 community screening measure of non-specific psychological distress.^20,21^ The measure has ‘substantial’ concordance with independent clinical ratings of serious mental illness.^21^ Participants were asked:

“During the past 30 days, about how often, if at all, did you feel… nervous; hopeless; restless or fidgety; so depressed that nothing could cheer you up; that everything was an effort; worthless?”

The answer options were presented in a randomised order and for each the respondent indicated one of the following: “All of the time (scored 4); Most of the time (3); Some of the time (2); A little of the time (1); None of the time (0)”

A sum score with a possible range from 0 to 24 was calculated. Based on previous research scores of 13 and higher were categorised as serious distress, scores between 5-12 as moderate and less than 5 as no/minimal distress.^22^

#### Smoking status

Smoking status was ascertained using responses to the following question:

“Which of the following best applies to you?”

Those who responded with “I smoke cigarettes (including hand rolled) every day” and “I smoke cigarettes (including hand rolled), but not every day” were categorised as current cigarette smokers.

Those who responded with “I smoke cigarettes (including hand rolled) every day”, “I smoke cigarettes (including hand rolled), but not every day” and “I have stopped smoking completely in the last year” were categorised as past-year smokers.

Those indicating that they do not smoke cigarettes, but do smoke tobacco of some kind (e.g. Pipe, cigar or shisha) were excluded from the analysis (n=138) because they do not include measures of dependence that are measured for cigarette.

#### Smoking and quitting behaviour

##### Cigarette addiction

Cigarette addiction was measured using the heaviness of smoking index (HSI).^23^ This HSI uses two questions from the Fagerström Test for Cigarette Dependence: time to first cigarette in the morning after waking and the number of cigarettes smoked per day. Those with a score >4 are considered to have high addiction, and those with <4 considered to have low/moderate addiction.

##### Motivation to stop smoking

Motivation to stop smoking was assessed using the Motivation To Stop Scale^24^, a single-item measure with seven response options representing increasing motivation to quit. Responses were collapsed into two variables reflecting high vs. low or no motivation to stop smoking.^24^

##### Quit attempts

Quit attempts in the past month was measured among past year smokers using the question “How many serious attempts to stop smoking have you made in the last 12 months?”, and if one or more attempts were reported: “How long ago did your most recent serious quit attempt start?”.

We distinguished those who attempted to quit up to 1 month ago versus those who made no quit attempt or attempted to quit more than 1 month before the interview but were not successful.

##### Socio-demographic characteristics

The socio-demographic variables age (categories 16-24, 25-34, 35-44, 45-54, 55-64, and ≥65 years), sex (categories women vs other), occupation-based social grade (AB (higher and intermediate managerial, administrative and professional), C1 (supervisory, clerical and junior managerial, administrative and professional), C2 (skilled manual workers), D (semi□skilled and unskilled manual workers) and E (state pensioners, casual and lowest□grade workers, unemployed)), region of England (government office region including nine categories: North East, North West, Yorkshire and the Humber, East Midlands, West Midlands, East of England, London, South East, South West), and the presence of children in the household were measured.

##### Time period

The variable for time period included four months of data (April-July) each from the years 2016, 2017 and 2020. In this study, data from 2016 and 2017 were collapsed together to form a new variable reflecting April-July 2016-2017. These time periods were chosen because questions related to mental health outcomes were not included in the surveys during 2018 and 2019, and were only re-added from April 2020. The comparison of the same four month time period in 2020 and 2016-2017 sought to account for potential seasonality in mental health disorders.^25,26^

### Sample selection

Overall, 19,960 (unweighted) adults aged 18+ were surveyed. Of these, 3,640 past-year (current and recent ex) smokers were asked the mental health questions. Those who exclusively smoked cigars and pipes (n = 138), did not complete the mental health questions or selected ‘I don’t know’ or ‘prefer not to say’ in response to any of them (n = 399), or had missing data on any of the other variables included in the present analysis were excluded. This left a final unweighted sample size for analysis of 2,972 past-year smokers of which 2,418 were current smokers.

### Statistical analysis

To address our first aim, weighted proportions (95% CIs) were used to describe the prevalence of past-month moderate and severe distress, respectively, during the period of April-July 2020, and April-July 2016-2017 among past-year smokers.

We constructed separate logistic regression models to assess changes in moderate and severe distress, respectively, among smokers (past-year and current) between the two time periods (April-July 2020 vs April-July 2016-2017 as referent) and age (six categories with 16-24 as referent) and the interaction terms.

All associations are reported as odds ratios (ORs) with 95% confidence intervals (adjusted for sex, social grade and region). The inclusion of the time period*age interaction allowed us to examine psychological distress at different levels of age, which is of interest given the strong age gradient in risk of death from COVID-19.^17^

To address our second aim, we: i) calculated weighted proportions and chi-square statistics to compare the distribution of moderate and severe distress, respectively, within socio-demographic (age, sex, social grade, whether there were children in the house) and smoking characteristics (cigarette addiction, quit attempts and motivation to stop smoking) of past-year smokers during April-July 2020 and April-July 2016-2017; and ii) constructed a series of stratified logistic regression models to examine any changes within these socio-demographic and smoking characteristic sub-groups between April-July 2016-2017 (referent) and April-July 2020. All associations are reported as odds ratios (ORs) with 95% confidence intervals (adjusted for age, social grade, sex and region except where the covariate was the variable of interest).

Analysis was carried out in R version 3.6.0 in September 2020. A 2-sided P□<□.05 was considered statistically significant. The analysis plan was pre-registered online at https://osf.io/eh6sk/.

#### Sensitivity analysis

The same analyses reported for the primary analysis was conducted but comparing April-July 2020 with **all** months in 2016-2017.

#### Unregistered post-hoc analyses

We conducted further logistic regression models to explore changes in moderate and severe distress among recent ex-smokers (quit within the past year) between the two time periods (April-July 2020 vs April-July 2016-2017 as referent) and age (six categories with 16-24 as referent). We calculated Bayes factors (BF) for non-significant associations to explore whether they provided evidence for no effect (BF < 1/3) when compared to the alternative hypothesis or indicated data insensitivity (BF ≥ 1/3 and < 3).^27^ The alternative hypothesis was modelled using a half-normal distribution centred on zero, with a standard deviation equal to the expected effect size.

In 2016-2017 mental health data was only collected among current and recent ex-smokers. From April 2020 all respondents were asked questions about their mental health, allowing us to examine levels of distress across all categories of smoking status. Analysing distress according to smoking status contextualises the findings among smokers against those who have never smoked or have been abstinent for a long time. Further analyses were run exploring differences in the prevalence distress between April-July 2020 according to smoking status. The results are presented as weighted proportions (with 95% CIs). Logistic regression was used to estimate the association between distress and smoking status (four categories: Smoker, Stopped in the past year, Stopped >1 year ago and Never Smoker (referent)).

## RESULTS

A weighted total of 3,211 past-year smokers (mean (SD) age = 43.26 (17.11) years; 48.08% women) completed the STS survey between April-July in 2016 (n = 1,106), 2017 (n = 1,066) and 2020 (n = 1,039). Among the overall sample 748 (23.29%) reported moderate distress, and 293 (9.12%) reported severe distress. See Table 1 for an overview of the sample characteristics. Weighted prevalence statistics for moderate and severe distress among past-year smokers in 2016, 2017 and 2020 are shown in Figure 1.

**Table 1:** Characteristics of past-year smokers (weighted data).

| Characteristic | Total (n %) | Year |  |  |
| --- | --- | --- | --- | --- |
|  |  | 2016 (%) | 2017 (%) | 2020 (%) |
| **Year** |  |  |  |  |
| 2016 | 1106 (34.44) | - | - | - |
| 2017 | 1066 (33.20) | - | - | - |
| 2020 | 1039 (32.36) | - | - | - |
| **Past-month distress** |  |  |  |  |
| None | 2170 (67.58) | 786 (71.05) | 759 (71.19) | 625 (60.17) |
| Moderate | 748 (23.29) | 234 (21.16) | 215 (20.14) | 299 (28.79) |
| Severe | 293 (9.12) | 86 (7.79) | 92 (8.68) | 115 (11.04) |
| **Age** |  |  |  |  |
| 18-24 | 556 (17.32) | 189 (17.11) | 187 (17.52) | 180 (17.36) |
| 25-34 | 822 (25.60) | 265 (23.99) | 238 (22.32) | 318 (30.64) |
| 35-44 | 579 (18.03) | 208 (18.78) | 198 (18.61) | 173 (16.60) |
| 45-54 | 545 (16.97) | 190 (17.21) | 211 (19.79) | 143 (13.80) |
| 55-64 | 376 (11.71) | 129 (11.67) | 125 (11.68) | 122 (11.77) |
| 65+ | 334 (10.40) | 282 (11.23) | 42 (10.07) | 12 (9.83) |
| **Social grade** |  |  |  |  |
| AB | 481 (14.98) | 164 (14.86) | 146 (13.68) | 171 (16.44) |
| C1 | 765 (23.82) | 265 (23.97) | 265 (24.89) | 235 (22.60) |
| C2 | 780 (24.29) | 288 (26.09) | 254 (23.80) | 238 (22.89) |
| D | 681 (21.21) | 214 (19.36) | 225 (21.09) | 242 (23.28) |
| E | 504 (15.70) | 174 (15.72) | 176 (16.53) | 154 (14.78) |
| **Sex** |  |  |  |  |
| Women | 1544 (48.08) | 509 (46.00) | 536 (49.74) | 535 (48.56) |
| **Children in household** |  |  |  |  |
| Yes | 1143 (35.60) | 407 (36.85) | 371 (34.77) | 365 (35.07) |
| No | 2068 (64.40) | 698 (63.15) | 695 (65.23) | 675 (64.93) |

Unweighted n = 2,972. \*Year = 4 month time period (April-July) of specified year. Social grade: AB = Higher managerial, administrative and professional; B =Intermediate managerial, administrative and professional; C1 = Supervisory, clerical and junior managerial, administrative and professional; C2 = Skilled manual workers; D = Semi-skilled and unskilled manual workers; E = State pensioners, casual and lowest grade workers, unemployed with state benefits only; Other = responses of "Men" or "In another way"

**Figure 1:**
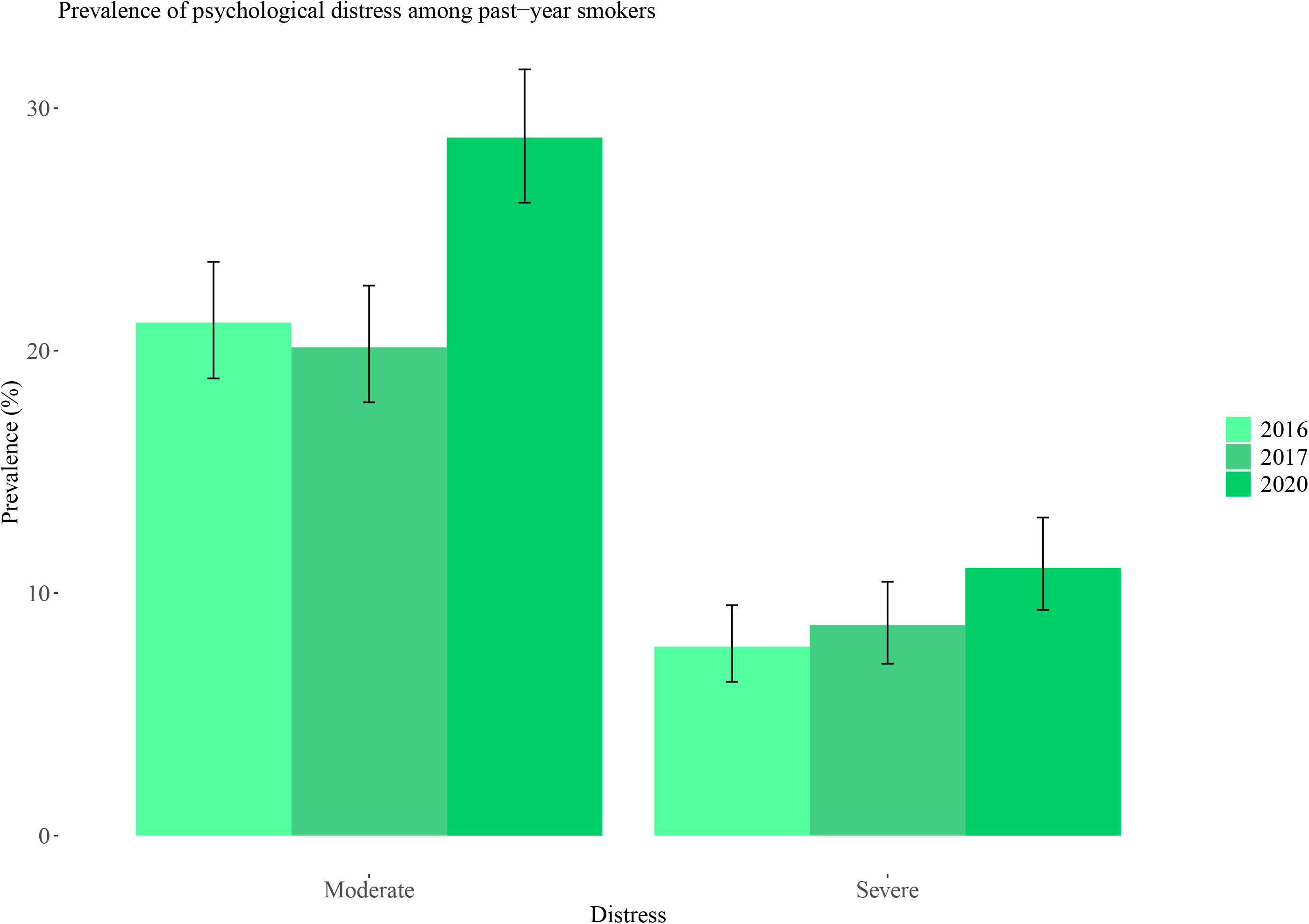
Prevalence of psychological distress among past-year smokers (weighted data)

### Changes in distress between 2016-2017 and 2020

#### Past-year smokers

Past-year smokers in 2020 had twice the odds of moderate and severe distress, respectively, compared with 2016-2017 (Table 2). An age gradient was apparent, with older age groups less likely to report moderate or severe distress, respectively, compared with those aged 16-24. There was no evidence of an interaction between time period and age.

**Table 2:** Associations between i) moderate (vs none) and ii) severe psychological distress (vs none) and time period of survey (April-July 2020 vs April-July 2016-2017) among past-year and current smokers in England

|  | Past-year smokers |  |  |  | Current smokers |  |  |  |
| --- | --- | --- | --- | --- | --- | --- | --- | --- |
|  | Moderate distress<sup>a</sup><br>(n=2,706) | *P* | Severe distress<sup>b</sup><br>(n=2,293) | *P* | Moderate distress<sup>c</sup><br>(n=2,455) | *P* | Severe distress<sup>d</sup><br>(n=2,095) | *P* |
| **Time period** |  |  |  |  |  |  |  |  |
| 2016-2017 ref | 1 [Reference] |  | 1 [Reference] |  | 1 [Reference] |  | 1 [Reference] |  |
| 2020 | 2.08 (1.34-3.25) | **.001** | 2.16 (1.13-4.07) | **.02** | 2.14 (1.31-3.50) | **.002** | 1.99 (0.95-4.01) | .06 |
| **Age** |  |  |  |  |  |  |  |  |
| 16-25 | 1 [Reference] |  | 1 [Reference] |  | 1 [Reference] |  | 1 [Reference] |  |
| 25-34 | 0.67 (0.48-0.94) | **.02** | 0.60 (0.36-0.98) | **.04** | 0.63 (0.44-0.89) | **.01** | 0.57 (0.33-0.96) | **.03** |
| 35-44 | 0.67 (0.47-0.95) | **.03** | 0.68 (0.4-1.13) | .14 | 0.67 (0.46-0.97) | **.03** | 0.75 (0.44-1.26) | 0.28 |
| 45-54 | 0.46 (0.32-0.66) | **<.001** | 0.54 (0.31-0.90) | **.02** | 0.46 (0.32-0.68) | **<.001** | 0.5 (0.28-0.86) | **.01** |
| 55-64 | 0.46 (0.31-0.68) | **<.001** | 0.51 (0.29-0.88) | **.02** | 0.49 (0.32-0.72) | **<.001** | 0.45 (0.24-0.8) | **.01** |
| 65+ | 0.22 (0.14-0.34) | **<.001** | 0.12 (0.05-0.25) | **<.001** | 0.23 (0.14-0.36) | **<.001** | 0.12 (0.05-0.26) | **<.001** |
| **Interaction terms** |  |  |  |  |  |  |  |  |
| 2020*25-34 | 0.79 (0.44-1.41) | .42 | 0.91 (0.39-2.13) | .82 | 0.78 (0.41-1.48) | .45 | 1.03 (0.41-2.64) | .95 |
| 2020*35-44 | 0.76 (0.39-1.45) | .40 | 0.78 (0.31-1.99) | .60 | 0.85 (0.42-1.73) | .66 | 0.87 (0.32-2.4) | .79 |
| 2020*45-54 | 0.95 (0.49-1.81) | .87 | 0.55 (0.19-1.49) | .25 | 0.93 (0.46-1.87) | .83 | 0.62 (0.20-1.84) | .39 |
| 2020*55-64 | 0.59 (0.3-1.17) | .13 | 0.39 (0.13-1.11) | .08 | 0.51 (0.24-1.06) | .07 | 0.55 (0.18-1.68) | .30 |
| 2020*65+ | 0.82 (0.38-1.75) | .61 | 1.31 (0.38-4.65) | .67 | 0.74 (0.32-1.66) | .45 | 1.07 (0.27-4.20) | .92 |

Ns are not weighted. All models are adjusted for age, sex and region. <sup>a</sup>Sample includes past-year smokers with moderate (n=679) and none/minimal (n=2,027) distress; <sup>b</sup>Sample includes past-year smokers with severe (n=266) and none/minimal (n=2,027) distress. <sup>c</sup>Sample includes current smokers with moderate (n=600) and none/minimal (n=1,855) distress; <sup>d</sup>Sample includes current smokers with severe (n=240) and none/minimal (n=1,855) distress. Models are adjusted for social grade, sex and region.

A sensitivity analysis using the entire two-year period of 2016 and 2017 as a comparator time period produced similar results to the main analysis. (Supplementary Table s1).

#### Current smokers

Similarly, current smokers in 2020 had twice the odds of moderate distress compared with 2016-2017 (Table 2). An age gradient was also apparent among current smokers, with older age groups less likely to report moderate or severe distress, respectively, compared with those aged 16-24. There was no evidence of an interaction between time period and age.

#### Recent ex-smokers

Among the much smaller group of recent ex-smokers (n=277) there were no significant associations between moderate or severe psychological distress, respectively, in 2020 compared with 2016-2017 (Supplementary Table s2). Based on observed increases in mental health problems pre and post COVID-19 among the general population in the UK^28^, exploratory expected effect sizes (ORs) were set to 1.1, 1.5 and 1.9 respectively. The calculation of Bayes factors under all of these contexts indicated that the data were insensitive to detect these effects (Supplementary Table s3).

#### Prevalence of psychological distress according to smoking status between April-July 2020

There were greater levels of both moderate and severe distress among smokers, recent and >1year ex-smokers compared with never smokers (Supplementary Figure s1 and Table s4).

### The distribution of distress within sociodemographic and smoking characteristics of past-year smokers in 2016-2017 and 2020

#### Moderate distress

The prevalence of moderate distress was higher in 2020 compared with 2016-2017 among: those aged 16-24 and 45-54, women, those in more disadvantaged social grades, those with and without children in the house, and among those with low cigarette addiction (Table 3). No differences were apparent among those who had tried to quit within the past month or among current smokers with high motivation to quit.

**Table 3:** Changes in the sociodemographic profile of i) no/minimal, ii) moderate and iii) severe distress in 2016/2017 and 2020.

| | 2016-2017<br>n (%) | 2020<br>n (%) | $\chi^2$ | P | ORadj (95%<br>CI) | P |
| --- | --- | --- | --- | --- | --- | --- |
| **No/minimal distress** | 1471 (70.93) | 556 (61.92) | - | - | - | - |
| **Age** |  |  |  |  |  |  |
| 16-24 | 225 (58.59) | 59 (43.70) | **8.47** | **.004** | **0.47 (0.3-0.72)** | **<.001** |
| 25-34 | 278 (67.97) | 132 (57.89) | **6.05** | **.01** | **0.63 (0.44-0.91)** | **.01** |
| 35-44 | 232 (68.24) | 77 (58.78) | **3.34** | **.06** | **0.58 (0.37-0.91)** | **.02** |
| 45-54 | 271 (74.86) | 90 (64.75) | **4.61** | **.03** | **0.48 (0.3-0.77)** | **.003** |
| 55-64 | 208 (73.50) | 104 (72.73) | <.001 | .96 | 0.93 (0.56-1.54) | .78 |
| 65+ | 257 (86.82) | 94 (77.05) | **5.43** | **.02** | **0.47 (0.26-0.84)** | **.01** |
| **Sex** |  |  |  |  |  |  |
| Women | 669 ( 66.57) | 256 ( 54.12) | **20.74** | **<.001** | **0.48 (0.38-0.61)** | **<.001** |
| Other* | 802 ( 75.02) | 300 ( 70.59) | 2.87 | .09 | **0.76 (0.59-0.99)** | **.04** |
| **Social grade** |  |  |  |  |  |  |
| AB | 216 (77.42) | 116 (78.91) | 0.05 | .82 | 1.08 (0.64-1.84) | .77 |
| C1 | 444 (75.00) | 177 (61.03) | **17.56** | **<.001** | **0.41 (0.29-0.57)** | **<.001** |
| C2 | 344 (76.79) | 125 (62.81) | **12.80** | **<.001** | **0.51 (0.35-0.75)** | **<.001** |
| D | 279 (71.91) | 75 (52.08) | **17.66** | **<.001** | **0.43 (0.28-0.65)** | **<.001** |
| E | 188 (51.23) | 63 (53.39) | 0.09 | .76 | 0.90 (0.57-1.41) | .64 |
| **Children in house** |  |  |  |  |  |  |
| Yes | 480 ( 70.18) | 177 (62.32) | **5.32** | **.02** | **0.6 (0.44-0.82)** | **.001** |
| No | 991 ( 71.29) | 379 (61.73) | **17.59** | **<.001** | **0.58 (0.47-0.73)** | **<.001** |
| **HSI** |  |  |  |  |  |  |
| Low (<4) | 1319 (72.31) | 508 (62.48) | **25.07** | **<.001** | **0.61 (0.51-0.73)** | **<.001** |
| High (≥4) | 152 (60.80) | 48 (56.47) | 0.33 | .57 | 0.67 (0.38-1.16) | .15 |
| **Quit attempt** |  |  |  |  |  |  |
| In past month | 85 (68.00) | 21 ( 63.64) | 0.07 | .79 | 0.68 (0.29-1.62) | .37 |
| **MTSS\*** |  |  |  |  |  |  |
| In ≤ 3 months | 190 (67.86) | 84 (70.59) | 0.18 | .67 | 1.07 (0.66-1.76) | .78 |
| **Moderate distress** | 429 (20.68) | 250 (27.84) | - | - | - | - |
| **Age** |  |  |  |  |  |  |
| 16-24 | 112 (21.17) | 56 (41.48) | **6.37** | **.01** | **2.05 (1.32-3.18)** | **.001** |
| 25-34 | 96 (23.47) | 68 (29.82) | **2.77** | **.10** | 1.37 (0.93-2) | .11 |
| 35-44 | 76 (22.35) | 37 (28.24) | 1.49 | .22 | 1.44 (0.89-2.30) | .13 |
| 45-54 | 62 (17.13) | 39 (28.06) | **6.79** | **.01** | **2.13 (1.3-3.48)** | **.003** |
| 55-64 | 51 (18.02) | 30 (20.98) | 0.36 | .55 | 1.17 (0.67-2.00) | .58 |
| 65+ | 32 (10.81) | 20 (16.39) | 1.99 | .16 | 1.74 (0.9-3.29) | .09 |
| **Sex** |  |  |  |  |  |  |
| Women | 229 ( 22.79) | 156 ( 32.98) | **16.83** | **<.001** | **1.85 (1.44-2.38)** | **<.001** |
| Other | 200 ( 18.71) | 94 (22.12) | 2.02 | .15 | 1.27 (0.95-1.69) | .10 |
| **Social grade** |  |  |  |  |  |  |
| AB | 50 (17.92) | 21 ( 14.29) | 0.67 | .41 | 0.74 (0.4-1.32) | 0.32 |
| C1 | 111 (18.75) | 89 ( 30.69) | **15.15** | **<.001** | **2.31 (1.63-3.29)** | **<.001** |
| C2 | 80 (17.86) | 57 ( 28.64) | **8.97** | **.003** | **1.86 (1.23-2.79)** | **.003** |
| D | 75 (19.33) | 50 (34.72) | **13.00** | **<.001** | **2.10 (1.35-3.26)** | **<.001** |
| E | 113 (30.79) | 33 (27.97) | 0.22 | .64 | 1.03 (0.63-1.65) | .92 |
| **Children in house** |  |  |  |  |  |  |
| Yes | 138 (20.18) | 76 (26.76) | **4.68** | **.03** | **1.57 (1.12-2.20)** | **.01** |
| No | 291 (20.94) | 174 (28.34) | **12.69** | **<.001** | **1.60 (1.27-2.02)** | **<.001** |
| **HSI** |  |  |  |  |  |  |
| Low (<4) | 368 (20.18) | 227 (27.92) | **18.87** | **<.001** | **1.57 (1.29-1.91)** | **<.001** |
| High (≥4) | 61 (24.40) | 23 (27.06) | 0.12 | .73 | 1.24 (0.67-2.24) | .48 |
| **Quit attempt** |  |  |  |  |  |  |
| In past month | 31 ( 24.80) | 7 (21.21) | 0.04 | .84 | 0.98 (0.34-2.59) | .98 |
| **MTSS\*** |  |  |  |  |  |  |
| In ≤ 3 months | 68 (24.29) | 26 (21.85) | 0.16 | .69 | 0.91 (0.53-1.51) | .71 |
| **Severe distress** | 174 (8.39) | 92 (10.24) | - | - | - | - |
| **Age** |  |  |  |  |  |  |
| 16-24 | 47 (12.24) | 20 (14.81) | 0.38 | .54 | 1.55 (0.89-2.71) | .12 |
| 25-34 | 35 (8.56) | 28 (12.28) | 1.88 | .17 | 1.22 (0.66-2.21) | .52 |
| 35-44 | 32 (9.41) | 17 (12.98) | 0.94 | .33 | 1.73 (0.86-3.43) | .12 |
| 45-54 | 29 (8.01) | 10 (7.19) | 0.01 | .90 | 1.23 (0.52-2.74) | .62 |
| 55-64 | 24 (8.48) | 9 (6.29) | 0.37 | .55 | 0.85 (0.34-2.00) | .72 |
| 65+ | 7 (2.36) | 8 (6.56) | 3.26 | .07 | **3.31 (1.04-10.95)** | **.04** |
| **Sex** |  |  |  |  |  |  |
| Women | 107 ( 10.65) | 61 (12.90) | 1.40 | .24 | **1.52 (1.07-2.16)** | **.02** |
| Other | 67 (6.27) | 31 ( 7.29) | 0.37 | .54 | 1.24 (0.78-1.95) | .36 |
| **Social grade** |  |  |  |  |  |  |
| AB | 13 (4.66) | 10 (6.80) | 0.50 | .48 | 1.68 (0.67-4.13) | 0.26 |
| C1 | 37 (6.25) | 24 (8.28) | 0.95 | .33 | 1.52 (0.84-2.7) | .16 |
| C2 | 24 (5.36) | 17 (8.54) | 1.85 | .17 | 1.52 (0.76-2.97) | .22 |
| D | 34 (8.76) | 19 (13.19) | 1.83 | .18 | 1.63 (0.86-3.06) | .13 |
| E | 66 (17.98) | 22 (18.64) | .001 | .98 | 1.15 (0.65-2.00) | .63 |
| **Children in house** |  |  |  |  |  |  |
| Yes | 66 (9.65) | 31 (10.92) | 0.23 | .63 | 1.39 (0.85-2.22) | .18 |
| No | 108 (7.77) | 61 (9.93) | 2.31 | .13 | 1.38 (0.97-1.96) | .07 |
| **HSI** |  |  |  |  |  |  |
| Low (<4) | 137 (7.51) | 78 (9.59) | 2.99 | .08 | **1.38 (1.02-1.86)** | **.03** |
| High (≥4) | 37 (14.80) | 14 (16.47) | 0.04 | .85 | 1.47 (0.69-3.03) | .31 |
| **Quit attempt** |  |  |  |  |  |  |
| In past month | 9 (7.20) | 5 (15.15) | 1.17 | .28 | 2.91 (0.73-11.2), | .12 |
| **MTSS\*** |  |  |  |  |  |  |
| In ≤ 3 months | 22 (7.86) | 9 (7.56) | **<.001** | 1.00 | 1.06 (0.44-2.37) | .90 |

Ns are not weighted. HSI = heaviness of smoking index. \*MTSS = motivation to stop smoking. MTSS is measured among current cigarette smokers only. Models are adjusted for age, social grade, sex and region except where the covariate was the variable of interest. Other = responses of "Men" or "In another way"

#### Severe distress

The prevalence of severe distress was higher in 2020 compared with 2016-2017 among: those aged 65+, women, and among those with low cigarette addiction (Table 3). There were no apparent differences in the prevalence of distress according to high cigarette addiction, recent quit attempts or motivation to stop smoking.

## DISCUSSION

Between April-July 2016/17 and April-July 2020 in England there were increases in moderate and severe distress, respectively, among both past-year and current smokers. Older age groups were less likely to report symptoms compared with younger groups, but there was no interaction between age and time period. Within socio-demographic categories, moderate distress was greater in 2020 among those aged 16-24 and 45-54 years, women, those from more disadvantaged social grades, those with and without children at home and those with low cigarette addiction. Severe distress was greater in 2020 among those aged 65+, women and among those with low cigarette addiction.

The increase in levels of both moderate and severe distress among smokers is likely caused by the ongoing COVID-19 pandemic and associated stay-at-home restrictions that have affected work and social life in England.^29^ Moreover, while mental health has deteriorated in the overall population as a result of COVID-19^12^, our analysis using exclusively April-July 2020 data highlighted that smokers specifically continue to display elevated levels of distress compared with non-smokers. Together these findings have concerning implications for existing smoking-related health inequalities considering the strong and potentially bi-directional associations between smoking and mental illness.^8^

Older smokers were less likely to report distress compared with younger groups.^11^ These findings re-emphasise the need to address higher prevalence of poor mental health among younger smokers.^12,30,31^ We hypothesised that there may be an interaction between age-group and year with older smokers experiencing greater distress in 2020 due to the age gradient in deaths from COVID-19 and the known risks of smoking. This was not borne out in the primary analysis, but there were signals of age-group differences in our stratified socio-demographic analyses discussed below.

Compared with 2016/17 the distribution of moderate distress in 2020 differed within certain sociodemographic characteristics, with higher prevalence in those aged 16-24 and 45-54, women, those from more disadvantaged social grades and in both those with and without children in the home. The distribution of severe distress was broadly similar across the two time periods, with the exceptions of higher prevalence in 2020 within women and those aged 65+. These demographic profiles of distress have important implications for existing inequalities and support other findings that the impacts of COVID-19 on worsening mental health have not been felt equally across society, but specifically among women and the more socioeconomically disadvantaged.^12^ However, the reported significance of these stratified socio-demographic analyses within age specifically (given the absence of interaction effects described above), but also other characteristics should be treated with caution and viewed descriptively.

Regarding smoking and quitting behaviour, our results indicate that between 2016/17 and 2020 there have been no changes in the prevalence of high cigarette addiction among those with moderate or severe distress, respectively. There were, however, increases in the prevalence of low addiction. Whether this phenomenon is unique to 2020 or represents a sustained change in the mental health population (as has been observed in the general population of smokers in England^32^) will be monitored going forward. The prevalence of past month quit attempts or motivation to stop smoking (among current smokers) did not change between 2016/17 and 2020 within any category of psychological distress. This is consistent with recently published findings in the general population showing that COVID-19 triggered only a minority of quit attempts in England.^33,34^

Recent YouGov data in England has suggested that those with existing mental health problems may have been more likely to have quit successfully during the pandemic.^35^ However, robust analyses of this unexpected finding is needed. Moreover, the same YouGov data has suggested that smokers with poor mental health who did quit during the pandemic are smoking more and are less likely to quit as a result of COVID-19.

Findings from this study have implications for policy and practice. The levels of distress among disadvantaged social grades and women, and the persistence of poor mental health among younger smokers is concerning given that the prevalence of mental illness 2016/17 was already thought to be greater in these demographics than in previous years.^36^ It is important that support for smoking cessation is available to those with mental health problems during the pandemic and in its aftermath. Mental health practitioners should continue to monitor the smoking status of their patients, and offer referral to local authority stop smoking services where they can receive effective support for smoking cessation.^37^ Specific attention should be considered for smokers aged 65+ at this time, who are generally more dependent on cigarettes^38^ and from our analyses appear to have greater distress than in previous years. Advice on effective harm reduction alternatives such as electronic cigarettes should also be considered.^39,40^ In addition, clear public health messaging about the immediate health benefits of smoking cessation is necessary to counter the uncertainty that has emerged related to some evidence that smokers appear to be at reduced risk of SARS-Cov-2 infection.^41^

To our knowledge this study is the first to analyse psychological distress among smokers using data before and during the COVID-19 pandemic. However, it is limited by the use of cross-sectional survey data where smoking status is self-reported. Moreover, we did not have mental health data in 2018, 2019 and the first three months of 2020. While it is likely that some of the increase in mental health problems in 2020 reflect a continuation of a secular trend going back to 2008^36^, data from the opinions and lifestyle survey collected between 2018 to date highlight a clear deterioration in wellbeing among smokers following the onset of the pandemic in England.^42^

Future research should continue to monitor changes in distress among smokers throughout the ongoing pandemic and its aftermath. Greater understanding about the direction(s) of the relationship between smoking and mental illness will also help inform the best approach for reducing smoking levels in this vulnerable group.

In conclusion, comparing April-July 2016/17 with April-July 2020 in England there were increases in moderate and severe distress among smokers. Continued support and messaging for smoking cessation among those with poor mental health is ever more important during the ongoing COVID-19 pandemic and in its aftermath.

## Data Availability

Pre-registered analysis plan and statistical code are available at https://osf.io/eh6sk/

## Funding and acknowledgements

We are grateful to Cancer Research UK and the UK Prevention Research Partnership for funding the study. Authors are members of the UK Prevention Research Partnership, an initiative funded by UK Research and Innovation Councils, the Department of Health and Social Care (England), and the UK devolved administrations and leading health research charities.

## References

1. Public Health England. Health matters: smoking and mental health. Health matters. Published 2020. Accessed August 20, 2020. https://www.gov.uk/government/publications/health-matters-smoking-and-mental

2. Richardson S, McNeill A, Brose LS. Smoking and quitting behaviours by mental health conditions in Great Britain (1993–2014). Addict Behav. 2019;90:14–19. doi:10.1016/j.addbeh.2018.10.011

3. Tam J, Warner KE, Meza R. Smoking and the Reduced Life Expectancy of Individuals With Serious Mental Illness. Am J Prev Med. 2016;51(6):958–966. doi:10.1016/j.amepre.2016.06.007

4. Pratt LA. Characteristics of Adults with Serious Psychological Distress as Measured by the K6 Scale: United States, 2001-04. Vol 382.; 2001. Accessed August 20, 2020. https://stacks.cdc.gov/view/cdc/6840

5. Lawrence D, Mitrou F, Zubrick SR. Non-specific psychological distress, smoking status and smoking cessation: United States National Health Interview Survey 2005. BMC Public Health. 2011;11(1):1–13. doi:10.1186/1471-2458-11-256

6. Streck JM, Weinberger AH, Pacek LR, Gbedemah M, Goodwin RD. Cigarette smoking quit rates among persons with serious psychological distress in the united states from 2008 to 2016: Are mental health disparities in cigarette use increasing? Nicotine Tob Res. 2020;22(1):130–134. doi:10.1093/ntr/nty227

7. Kim-Mozeleski JE, Pandey R, Tsoh JY. Psychological distress and cigarette smoking among US households by income: Considering the role of food insecurity. Prev Med reports. 2019;16:100983.

8. Fluharty M, Taylor AE, Grabski M, Munafò MR. The association of cigarette smoking with depression and anxiety: A systematic review. Nicotine Tob Res. 2017;19(1):3–13. doi:10.1093/ntr/ntw140

9. Boden JM, Fergusson DM, Horwood LJ. Cigarette smoking and depression: tests of causal linkages using a longitudinal birth cohort. Br J Psychiatry. 2010;196(6):440–446.

10. Skov-Ettrup LS, Nordestgaard BG, Petersen CB, Tolstrup JS. Does high tobacco consumption cause psychological distress? A Mendelian randomization study. Nicotine Tob Res. 2016;19(1):32–38.

11. Brose LS, Brown J, Robson D, McNeill A. Mental health, smoking, harm reduction and quit attempts - A population survey in England. BMC Public Health. 2020;20(1):1237. doi:10.1186/s12889-020-09308-x

12. Pierce M, Hope H, Ford T, et al. Mental health before and during the COVID-19 pandemic: a longitudinal probability sample survey of the UK population. The Lancet Psychiatry. Published online 2020.

13. Boland VC, Mattick RP, McRobbie H, Siahpush M, Courtney RJ. “I’m not strong enough; I’m not good enough. i can’t do this, I’m failing”-A qualitative study of low-socioeconomic status smokers’ experiences with accesssing cessation support and the role for alternative technology-based support. Int J Equity Health. 2017;16(1):196. doi:10.1186/s12939-017-0689-5

14. Bishop S, Panjari M, Astbury J, et al. “A survey of antenatal clinic staff: some perceived barriers to the promotion of smoking cessation in pregnancy.” Aust Coll Midwives Inc J. 1998;11(3):14–18. http://ovidsp.ovid.com/ovidweb.cgi?T=JS&PAGE=reference&D=emed7&NEWS=N&AN=129348380

15. Sweet TM. A Review of Statistical Rethinking: A Bayesian Course With Examples in R and Stan. J Educ Behav Stat. 2017;42(1):107–110. doi:10.3102/1076998616659752

16. Jackson SE, Garnett C, Shahab L, Oldham M, Brown J. Association of the Covid□19 lockdown with smoking, drinking, and attempts to quit in England: an analysis of 2019□2020 data. Addiction. Published online October 21, 2020:add.15295. doi:10.1111/add.15295

17. Verity R, Okell LC, Dorigatti I, et al. Estimates of the severity of coronavirus disease 2019: a model-based analysis. Lancet Infect Dis. 2020;20(6):669–677. doi:10.1016/S1473-3099(20)30243-7

18. Fidler JA, Shahab L, West O, et al. “The smoking toolkit study”: a national study of smoking and smoking cessation in England. BMC Public Health. 2011;11:479. doi:10.1186/1471-2458-11-479

19. Knottnerus A, Tugwell P. STROBE--a checklist to Strengthen the Reporting of Observational Studies in Epidemiology. J Clin Epidemiol. 2008;61(4):323.

20. Kessler RC, Andrews G, Colpe LJ, et al. Short screening scales to monitor population prevalences and trends in non-specific psychological distress. Psychol Med. 2002;32(6):959–976.

21. Kessler RC, Green JG, Gruber MJ, et al. Screening for serious mental illness in the general population with the K6 screening scale: results from the WHO World Mental Health (WMH) survey initiative. Int J Methods Psychiatr Res. 2010;19(S1):4-22.

22. Prochaska JJ, Sung H, Max W, Shi Y, Ong M. Validity study of the K6 scale as a measure of moderate mental distress based on mental health treatment need and utilization. Int J Methods Psychiatr Res. 2012;21(2):88–97.

23. Kozlowski LT, Porter CQ, Orleans CT, Pope MA, Heatherton T. Predicting smoking cessation with self-reported measures of nicotine dependence: FTQ, FTND, and HSI. Drug Alcohol Depend. 1994;34(3):211–216.

24. Kotz D, Brown J, West R. Predictive validity of the Motivation To Stop Scale (MTSS): a single-item measure of motivation to stop smoking. Drug Alcohol Depend. 2013;128(1-2):15–19.

25. De Graaf R, Van Dorsselaer S, Ten Have M, Schoemaker C, Vollebergh WAM. Seasonal variations in mental disorders in the general population of a country with a maritime climate: Findings from the Netherlands mental health survey and incidence study. Am J Epidemiol. 2005;162(7):654–661. doi:10.1093/aje/kwi264

26. Magnusson A. An overview of epidemiological studies on seasonal affective disorder. Acta Psychiatr Scand. 2000;101(3):176–184. doi:10.1034/j.1600-0447.2000.101003176.x

27. Beard E, Dienes Z, Muirhead C, West R. Using Bayes factors for testing hypotheses about intervention effectiveness in addictions research. Addiction. 2016;111(12):2230–2247.

28. Daly M, Sutin A, Robinson E. Longitudinal changes in mental health and the COVID-19 pandemic: Evidence from the UK Household Longitudinal Study. Published online 2020.

29. Bu F, Steptoe A, Mak HW, Fancourt D. Time-use and mental health during the COVID-19 pandemic: a panel analysis of 55,204 adults followed across 11 weeks of lockdown in the UK. medRxiv. Published online August 21, 2020:2020.08.18.20177345. doi:10.1101/2020.08.18.20177345

30. Pedersen W, Von Soest T. Smoking, nicotine dependence and mental health among young adults: a 13□year population□based longitudinal study. Addiction. 2009;104(1):129–137.

31. Schmidt AM, Golden SD, Gottfredson NC, Ennett ST, Aiello AE, Ribisl KM. Psychological Health and Smoking in Young Adulthood. Emerg Adulthood. Published online 2019:2167696819858812.

32. Garnett C, Tombor I, Beard E, Jackson SE, West R, Brown J. Changes in smoker characteristics in England between 2008 and 2017. Addiction. 2020;115(4):748–756. doi:10.1111/add.14882

33. Heerfordt C, Heerfordt IM. Has there been an increased interest in smoking cessation during the first months of the COVID-19 pandemic? A Google Trends study. Public Health. 2020;183:6.

34. Tattan□Birch H, Perski O, Jackson S, Shahab L, West R, Brown J. COVID□19, smoking, vaping and quitting: a representative population survey in England. Addiction. Published online September 28, 2020:add.15251. doi:10.1111/add.15251

35. Action on Smoking and Health. Supporting smokers to quit can help tackle surge in poor mental health. Published 2020. Accessed August 20, 2020. https://ash.org.uk/media-and-news/press-releases-media-and-news/supporting-smokers-to-quit-can-help-tackle-surge-in-poor-mental-health/

36. Slee A, Nazareth I, Freemantle N, Background LH. Trends in generalised anxiety disorders and symptoms in primary care: UK population-based cohort study. doi:10.1192/bjp.2020.159

37. Gilbody S, Peckham E, Bailey D, et al. Smoking cessation for people with severe mental illness (SCIMITAR+): a pragmatic randomised controlled trial. The Lancet Psychiatry. 2019;6(5):379–390.

38. Hall SM, Humfleet GL, Gorecki JA, Muñoz RF, Reus VI, Prochaska JJ. Older versus younger treatment-seeking smokers: differences in smoking behavior, drug and alcohol use, and psychosocial and physical functioning. Nicotine Tob Res. 2008;10(3):463–470.

39. Hartmann_Boyce J, McRobbie H, Lindson N, et al. Electronic cigarettes for smoking cessation. Cochrane Database Syst Rev. 2020;(10). doi:10.1002/14651858.CD010216.pub4

40. Hickling LM, Perez-Iglesias R, McNeill A, et al. A pre-post pilot study of electronic cigarettes to reduce smoking in people with severe mental illness. Psychol Med. 2019;49(6):1033–1040.

41. Simons D, Shahab L, Brown J, Perski O. The association of smoking status with SARS□CoV□2 infection, hospitalisation and mortality from COVID□19: A living rapid evidence review with Bayesian meta□analyses (version 7). Addiction. Published online October 2, 2020:add.15276. doi:10.1111/add.15276

42. Public Health England. Wider Impacts of COVID-19. Wider Impacts of COVID-19 on Health (WICH) monitoring tool. Published 2020. Accessed October 30, 2020. https://analytics.phe.gov.uk/apps/covid-19-indirect-effects/

