## Supplementary Materials for "Smoking, distress and COVID-19 in England: cross-sectional population surveys from 2016 to 2020"

**SUPPLEMENTARY FILE**

**Table s1**: Associations between i) moderate and ii) severe psychological distress and time period of survey (April-July 2020 vs all months in 2016-2017) among past-year smokers in England

|  | **Past-year smokers** | | | |
| --- | --- | --- | --- | --- |
|  | **Moderate distress^a^** | ***P*** | **Severe distress^b^** | ***P*** |
|  | (n=6,501) |  | (n=5,524) |  |
| **Time period** |  |  |  |  |
| 2016-2017 ref | 1 [Reference] |  | 1 [Reference] |  |
| 2020 | 2.02 (1.36-3.01) | **<.001** | 2.10 (1.17-3.64) | **.01** |
| **Age** |  |  |  |  |
| 16-25 | 1 [Reference] |  | 1 [Reference] |  |
| 25-34 | 0.69 (0.57-0.83) | **<.001** | 0.71 (0.54-0.95) | **.02** |
| 35-44 | 0.72 (0.59-0.88) | **.001** | 0.64 (0.47-0.86) | **.004** |
| 45-54 | 0.53 (0.43-0.65) | **<.001** | 0.58 (0.43-0.78) | **<.001** |
| 55-64 | 0.41 (0.33-0.52) | **<.001** | 0.40 (0.28-0.56) | **<.001** |
| 65+ | 0.26 (0.2-0.33) | **<.001** | 0.10 (0.06-0.16) | **<.001** |
| **Interaction terms** |  |  |  |  |
| 2020*25-34 | 0.77 (0.46-1.28) | .31 | 0.79 (0.38-1.66) | .53 |
| 2020*35-44 | 0.68 (0.38-1.22) | .20 | 0.86 (0.38-1.96) | .73 |
| 2020*45-54 | 0.85 (0.48-1.5) | .57 | 0.52 (0.2-1.26) | .15 |
| 2020*55-64 | 0.70 (0.38-1.26) | .24 | 0.53 (0.2-1.31) | .18 |
| 2020*65+ | 0.73 (0.37-1.40) | .35 | 1.6 (0.54-4.49) | .38 |

Ns are not weighted. All models are adjusted for age, sex and region. ^a^Sample includes past-year smokers with moderate (n=1,593) and none/minimal (n=5,003) distress; ^b^Sample includes past-year smokers with severe (n=599) and none/minimal (n=5,003) distress. Models are adjusted for age, sex and region.

**Post-hoc analyses**

**Table s2**: Associations between i) moderate and ii) severe psychological distress and time period of survey (April-July 2020 vs April-July 2016-2017) among recent ex-smokers in England

|  | **Moderate distress^a^** | ***P*** | **Severe distress^b^** | ***P*** |
| --- | --- | --- | --- | --- |
|  | (n=251) |  | (n=198) |  |
| **Time period** |  |  |  |  |
| 2016-2017 ref | 1 [Reference] |  | 1 [Reference] |  |
| 2020 | 1.33 (0.73-2.42) | .39 | 1.28 (0.48-3.41) | .62 |
| **Age** |  |  |  |  |
| 16-25 | 1 [Reference] |  | 1 [Reference] |  |
| 25-34 | 0.85 (0.37-1.92) | .69 | 0.55 (0.16-1.91) | .35 |
| 35-44 | 0.43 (0.16-1.09) | .08 | 0.09 (0-0.58) | **.03** |
| 45-54 | 0.41 (0.15-1.11) | .08 | 0.41 (0.09-1.64) | .22 |
| 55-64 | 0.30 (0.09-0.90) | **.04** | 0.65 (0.14-2.6) | .55 |
| 65+ | 0.15 (0.04-0.49) | **.003** | 0.24 (0.03-1.13) | .10 |

Model adjusted for social grade, sex, and region

**Table s3**: Bayes factor calculation for associations between moderate and severe psychological distress and time period of survey among recent ex-smokers in England.

| **Moderate Distress** |  |
| --- | --- |
| **Expected effect size OR = 1.1** |  |
| Sample standard error | 0.30 |
| Obtained sample estimate | 0.29 |
| Mean of alternative hypothesis (half-normal) | 0 |
| Plausible expected value | 0.095 |
| Number of tails | 1 |
| Bayes factor | 1.21 |
| **Expected effect size OR = 1.5** |  |
| Sample standard error | 0.30 |
| Obtained sample estimate | 0.29 |
| Mean of alternative hypothesis (half-normal) | 0 |
| Plausible expected value | 0.41 |
| Number of tails | 1 |
| Bayes factor | 1.23 |
| **Expected effect size OR = 1.9** |  |
| Sample standard error | 0.30 |
| Obtained sample estimate | 0.29 |
| Mean of alternative hypothesis (half-normal) | 0 |
| Plausible expected value | 0.64 |
| Number of tails | 1 |
| Bayes factor | 0.98 |
| **Severe distress** |  |
| **Expected effect size OR = 1.1** |  |
| Sample standard error | 0.30 |
| Obtained sample estimate | 0.26 |
| Mean of alternative hypothesis (half-normal) | 0 |
| Plausible expected value | 0.095 |
| Number of tails | 1 |
| Bayes factor | 1.17 |
| **Expected effect size OR = 1.5** |  |
| Sample standard error | 0.30 |
| Obtained sample estimate | 0.26 |
| Mean of alternative hypothesis (half-normal) | 0 |
| Plausible expected value | 0.41 |
| Number of tails | 1 |
| Bayes factor | 1.10 |
| **Expected effect size OR = 1.9** |  |
| Sample standard error | 0.30 |
| Obtained sample estimate | 0.26 |
| Mean of alternative hypothesis (half-normal) | 0 |
| Plausible expected value | 0.64 |
| Number of tails | 1 |
| Bayes factor | 0.86 |

**Figure s1:** Prevalence of psychological distress by smoking status in 2020

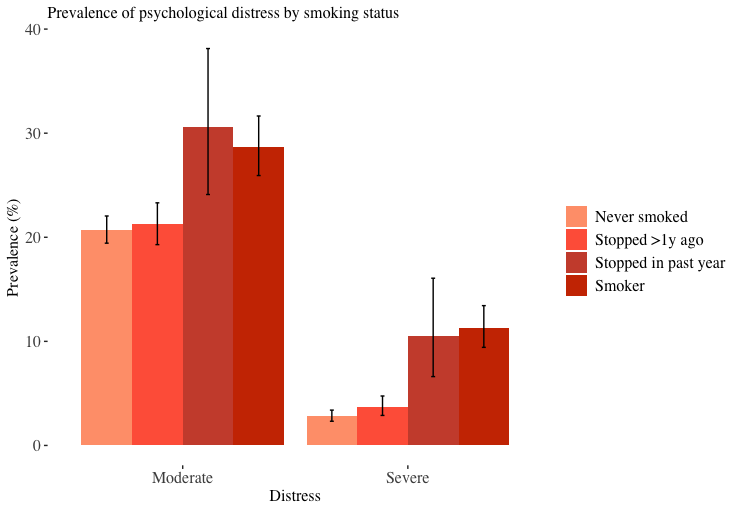

**Table s4**: Association between i) moderate and ii) severe psychological distress and smoking status between April-July 2020.

|  | **Moderate distress^a^** | ***P*** | **Severe distress^b^** | ***P*** |
| --- | --- | --- | --- | --- |
|  | (n=6192) |  | (n=5091) |  |
| **Smoking status** |  |  |  |  |
| Never smoked | 1 [Reference] |  | 1 [Reference] |  |
| Stopped >1y ago | 1.25 (1.07-1.46) | **.004** | 1.71 (1.19-2.43) | **.003** |
| Stopped past year | 1.61 (1.08-2.36) | **.02** | 4.84 (2.49-8.87) | **<.001** |
| Smoker | 1.46 (1.21-1.75) | **<.001** | 3.69 (2.64-5.15) | **<.001** |

Model is adjusted for age, sex, social grade and region.
